# Association between spironolactone use and COVID-19 outcomes in population-scale claims data: a retrospective cohort study

**DOI:** 10.1101/2023.02.28.23286515

**Authors:** Henry C. Cousins, Russ B. Altman

**Author notes:** **Corresponding author:** Russ B. Altman, M.D., Ph.D., 443 Via Ortega, MC 4245, Stanford, CA 94305.

## Abstract

**Background:** Spironolactone has been proposed as a potential modulator of SARS-CoV-2 cellular entry. We aimed to measure the effect of spironolactone use on the risk of adverse outcomes following COVID-19 hospitalization.

**Methods:** We performed a retrospective cohort study of COVID-19 outcomes for patients with or without exposure to spironolactone, using population-scale claims data from the Komodo Healthcare Map. We identified all patients with a hospital admission for COVID-19 in the study window, defining treatment status based on spironolactone prescription orders. The primary outcomes were progression to respiratory ventilation or mortality during the hospitalization. Odds ratios (OR) were estimated following either 1:1 propensity score matching (PSM) or multivariable regression. Subgroup analysis was performed based on age, gender, body mass index (BMI), and dominant SARS-CoV-2 variant.

**Findings:** Among 898,303 eligible patients with a COVID-19-related hospitalization, 16,324 patients (1.8%) had a spironolactone prescription prior to hospitalization. 59,937 patients (6.7%) met the ventilation endpoint, and 26,515 patients (3.0%) met the mortality endpoint. Spironolactone use was associated with a significant reduction in odds of both ventilation (OR 0.82; 95% CI: 0.75-0.88; p < 0.001) and mortality (OR 0.88; 95% CI: 0.78-0.99; p = 0.033) in the PSM analysis, supported by the regression analysis. Spironolactone use was associated with significantly reduced odds of ventilation for all age groups, men, women, and non-obese patients, with the greatest protective effects in younger patients, men, and non-obese patients.

**Interpretation:** Spironolactone use was associated with a protective effect against ventilation and mortality following COVID-19 infection, amounting to up to 64% of the protective effect of vaccination against ventilation and consistent with an androgen-dependent mechanism. The findings warrant initiation of large-scale randomized controlled trials to establish a potential therapeutic role for spironolactone in COVID-19 patients.

## INTRODUCTION

The continued proliferation of vaccine-evading severe acute respiratory syndrome coronavirus 2 (SARS-CoV-2) strains has reinforced the need for outpatient treatments to mitigate the clinical course of coronavirus disease 2019 (COVID-19).^1^ While a small number of antiviral therapies have received Food and Drug Administration approval in COVID-19, such treatments remain limited in both adoption and efficacy, owing to concerns about adverse reactions, drug-drug interactions, and cost.^2,3^ Consequently, it remains critical to identify any existing medications that may modulate the course of infection.

The potassium-sparing diuretic spironolactone has been proposed as a potential modulator of SARS-CoV-2 infection due to its interactions with multiple COVID-19-associated signaling pathways.^4^ Spironolactone functions chiefly as a mineralocorticoid receptor blocker, antagonizing the final stage of the renin-angiotensin-aldosterone system (RAAS).^5^ Given the involvement of angiotensin-converting enzyme 2 (ACE2), the canonical host receptor for SARS-CoV-2, in RAAS activity, mineralocorticoid antagonists have been hypothesized to alter ACE2 expression, which has been observed *in vitro*.^6,7^ In addition to its anti-mineralocorticoid effects, spironolactone is a strong inhibitor of the androgen receptor.^8^ The critical role of androgen signaling in upregulating *TMPRSS2*, which facilitates Spike processing during membrane fusion, suggests that spironolactone’s anti-androgenic activity could likewise impede viral entry.^9^

Existing clinical evidence for a protective role of spironolactone in COVID-19 is encouraging but inconclusive. One case-control study of 6,462 patients with liver cirrhosis in South Korea revealed a significant negative association between spironolactone use and COVID-19 diagnosis.^10^ A non-randomized, comparative study of bromhexine-spironolactone combination therapy in 103 patients identified a statistically significant 13% reduction in hospitalization time for the intervention group.^11^ The only published randomized, controlled clinical trial of spironolactone in COVID-19, to our knowledge, was a trial of sitagliptin-spironolactone combination therapy in 263 patients, which suggested a potentially beneficial effect for the intervention group with respect to clinical progression score.^12^

To determine whether spironolactone use is associated with COVID-19 severity, we conducted the largest clinical investigation of spironolactone in COVID-19 to date. Using health insurance claims data from public and private payers covering over 325 million unique patients, we performed a retrospective cohort study of COVID-19 outcomes for spironolactone users.

## METHODS

### Study design and population

We conducted a retrospective cohort study based on deidentified medical and pharmaceutical data from the Komodo Healthcare Map, a collection of health insurance claims from public and private payers nationwide. The database contains claims data for approximately 325 million unique patients in the United States since October 1, 2015 and is closely aligned with the National Health Interview Survey population in terms of geography and demographics. The dataset encompassed medical claims, pharmacy claims, enrollment records, and mortality records.

We identified 909,531 patients in the database who experienced a hospitalization due to COVID-19 within the study window, spanning March 1, 2020 to June 3, 2022. In the event that a patient experienced multiple COVID-19-related hospitalizations, only the first encounter was considered. Patients under the age of 15 years were also excluded. Medical variables were defined using International Classification of Diseases, 10th Revision, Clinical Modification (ICD-10-CM) codes, procedures were defined using ICD-10 Procedure Coding System (ICD-10-PCS), Current Procedural Technology (CPT), and Healthcare Common Procedure Coding System (HCPCS) codes, and drug prescriptions were defined using National Drug Code (NDC) identifiers. The study followed the Strengthening the Reporting of Observational Studies in Epidemiology (STROBE) guidelines.

### Ethics committee oversight

The study was declared exempt from institutional review board (IRB) review by the Stanford University IRB.

### Exposures and outcomes

Exposure was defined as a prescription for spironolactone within a 180-day window prior to the COVID-19-related hospitalization claim date.^13,14^ Only paid prescriptions, implying patient receipt of the medication, were considered, and multi-ingredient drugs were not included. The primary study outcome was progression to ventilation, defined as a claim for a respiratory ventilation procedure during the COVID-19-related hospitalization. We also considered mortality as an additional endpoint, which was defined as death recorded within the period covered by the COVID-19-related hospitalization claim. Time-stationarity of outcome variables was measured by Pearson correlation between endpoint probability and month of admission, beginning April 2020.

### Study variables

The study design controlled for demographic, medical, and pharmaceutical covariates. Demographic information included age as a continuous variable and gender. Medical covariates included body mass index (BMI ≥30 kg/m^2^ or <30 kg/m^2^), myocardial infarction, congestive heart failure, peripheral vascular disease, dementia, chronic pulmonary disease, rheumatic disease, peptic ulcer disease, mild liver disease, moderate or severe liver disease, diabetes without chronic complications, diabetes with chronic complications, hemiplegia or paraplegia, renal disease, malignancy, metastatic solid tumor, and human immunodeficiency virus (HIV) or acquired immunodeficiency syndrome (AIDS). For each condition, corresponding ICD-10-CM codes were obtained from the updated Charlson Comorbidity Index definitions (**Supplementary Table S1**).^15^ Pharmaceutical covariates included a paid prescription within a 180-day window prior to the COVID-19-related hospitalization for atorvastatin, levothyroxine, metformin, lisinopril, amlodipine, metoprolol, albuterol, omeprazole, losartan, or gabapentin.

COVID-19 vaccination status was an additional covariate, defined as receipt of at least one dose of any COVID-19 vaccine prior to the COVID-19-related hospitalization.

Values are reported as median (interquartile range [IQR]) for continuous variables and frequency (percent) for categorical variables.

### Propensity score matching

We performed propensity score matching (PSM) to obtain matched pairs of drug-exposed and non-drug-exposed patients.^16^ Propensity scores were derived by fitting a logistic regression model with L2 regularization to predict drug exposure status using all study covariates, normalized to unity. Nearest-neighbor matching on propensity scores was performed without a caliper to generate 1:1 matched pairs. Covariate balance between drug-exposed and non-drug-exposed groups was assessed by calculating the standardized bias for each covariate, with a standardized bias of less than 0.1 considered balanced (**Supplementary Table S2**).^17^ Odds ratios (OR) between drug exposure and each outcome of interest, as well as corresponding 95% confidence intervals (95% CI) and p-values, were calculated using McNemar’s exact test.

### Regression model

We also fit a second model to control for covariates without matching. We fit an L1-regularized logistic regression (LR) model using the same explanatory variables as in the propensity score derivation, with the addition of drug exposure status. OR were calculated from coefficients of the fitted model, and confidence intervals and p-values for each OR were calculated from the corresponding t statistics.

### Subgroup and sensitivity analysis

We measured treatment effects in patient subgroups, grouping by male gender, female gender, obesity (BMI≥30 kg/m^2^), non-obesity (BMI<30 kg/m^2^), and age brackets (<60, 60-74, and ≥75 years). Additionally, we analyzed cases in time periods with predominance of specific variants in the US, including the Delta (July 1, 2021 to December 20, 2021) and Omicron (December 20, 2021 to June 3, 2022) strains.^18^ We ran additional sensitivity analyses considering alternate windows of drug exposure (90 days and 360 days).

### Computational resources

Bulk data queries were performed using Structured Query Language (SQL) in a Snowflake workspace (Snowflake Inc., Bozeman, MT). All statistical analyses were performed in a Python 3.10 environment using the scikit-learn (version 1.1.2), statsmodels (version 0.13.2), psmpy (version 0.3.5), NumPy (version 1.23.2), and pandas (version 1.4.3) packages.

## RESULTS

From the database, we identified 909,531 patients with a COVID-19-related hospitalization, of whom 11,228 patients were excluded due to age below the study minimum (n = 11,206) or missing information for gender (n = 22; **Figure 1**). Within the final study population of 898,303, the treatment group comprised 16,324 patients with a fulfilled prescription for spironolactone prior to hospitalization.

**Figure 1.**
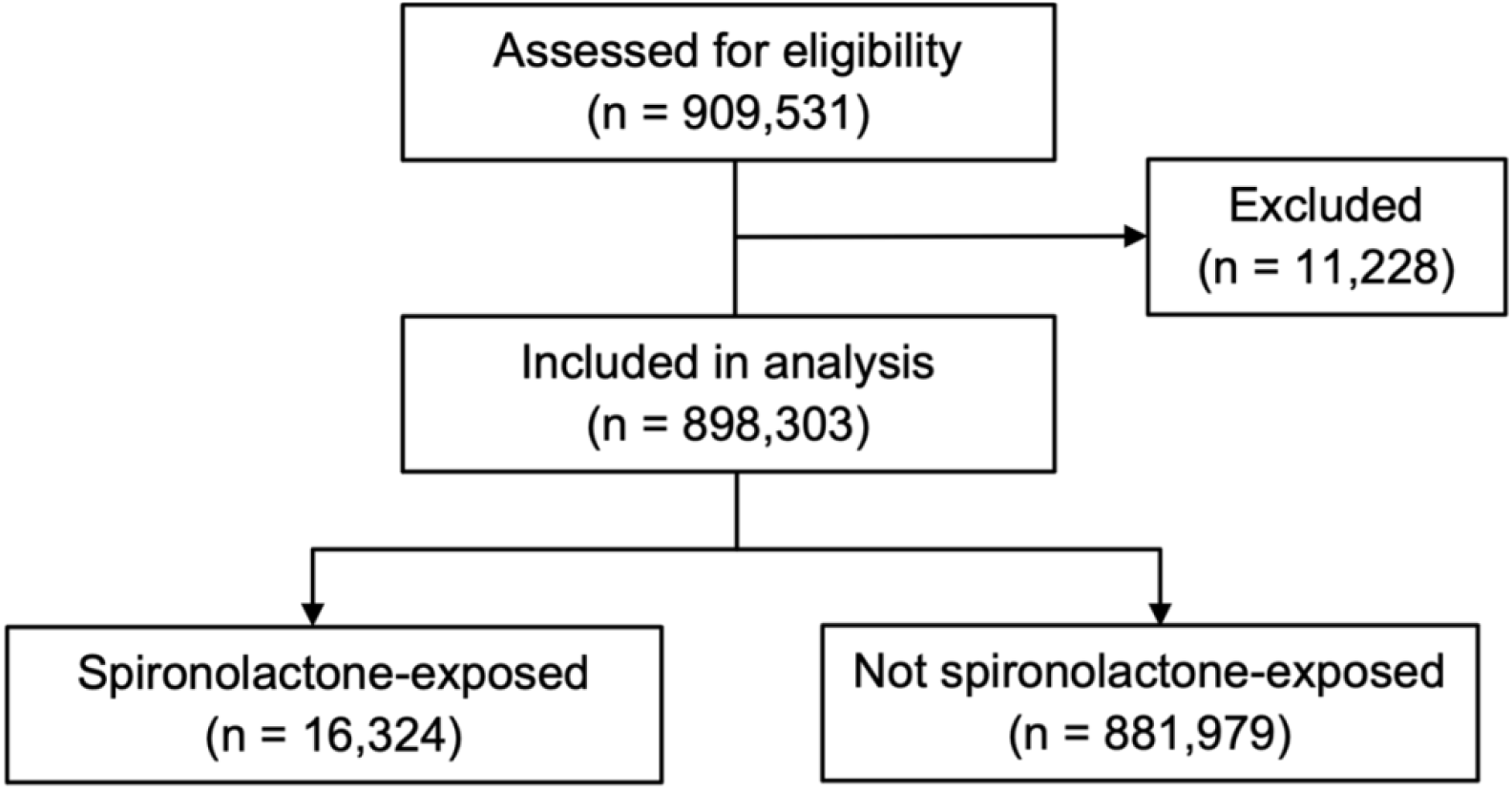
Selection of study population. 909,531 patients hospitalized for COVID-19 were identified from the claims database. 11,228 patients were excluded due to age below the study minimum (n = 11,206) or missing gender (n = 22). Of the 898,303 patients included in the analysis, 16,324 had a paid prescription for spironolactone within a 180-day window prior to the hospitalization.

The study population had a median age of 64.9 (IQR 56.5-78.6) years, and 465,124 (51.8%) were women. 59,937 patients (6.7%) met the ventilation endpoint, and 26,515 patients (3.0%) met the mortality endpoint. Endpoints were time-stationary, with no significant correlation between event frequency and claim month over the study window (p = 0.338 for ventilation; p = 0.248 for mortality). Following propensity score matching, all covariates were well balanced between treatment and control groups, with standardized biases of less than 0.1 in all cases (**Table I**).

**Table I.**
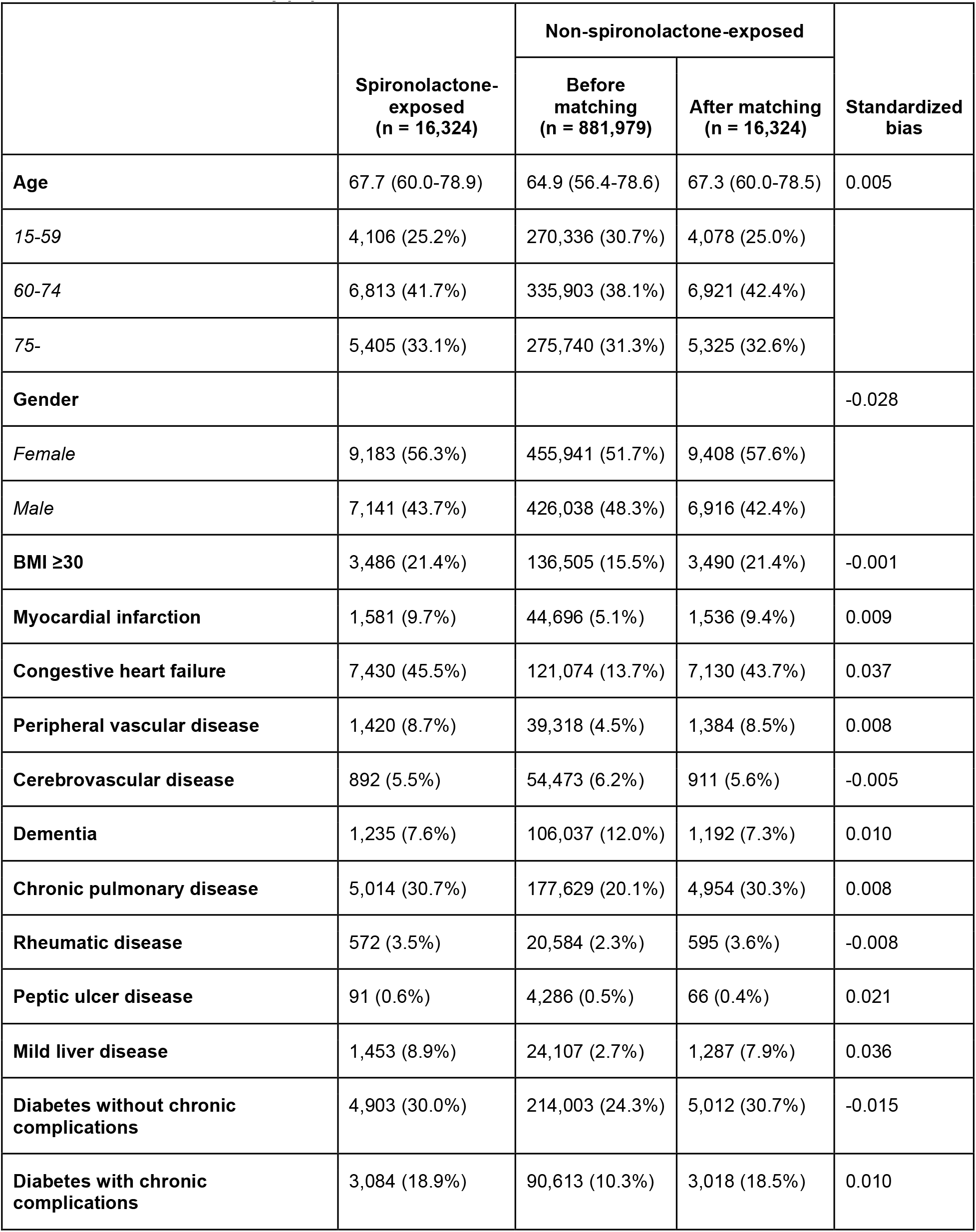
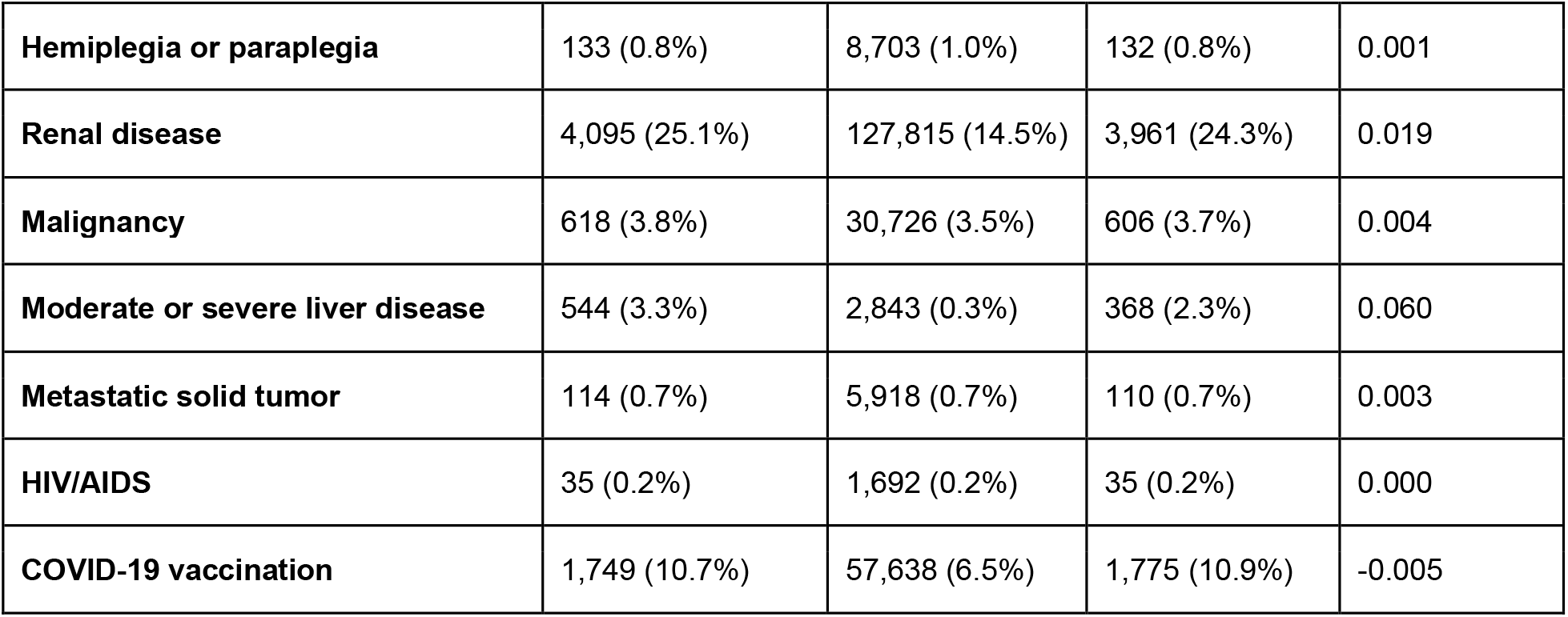
Characteristics of study population

Following 1:1 propensity score matching, 1,212 of 16,324 patients (7.4%) in the spironolactone treatment group met the ventilation endpoint in aggregate, while 1,459 of 16,324 patients (8.9%) in the control group met the endpoint (**Table II**). In the paired analysis, the OR for ventilation between treatment and controls was 0.82 (95% CI: 0.75-0.88; p < 0.001). The unmatched logistic regression analysis supported this protective effect, finding an OR of 0.78 (95% CI: 0.73-0.83; p < 0.001).

**Table II.**
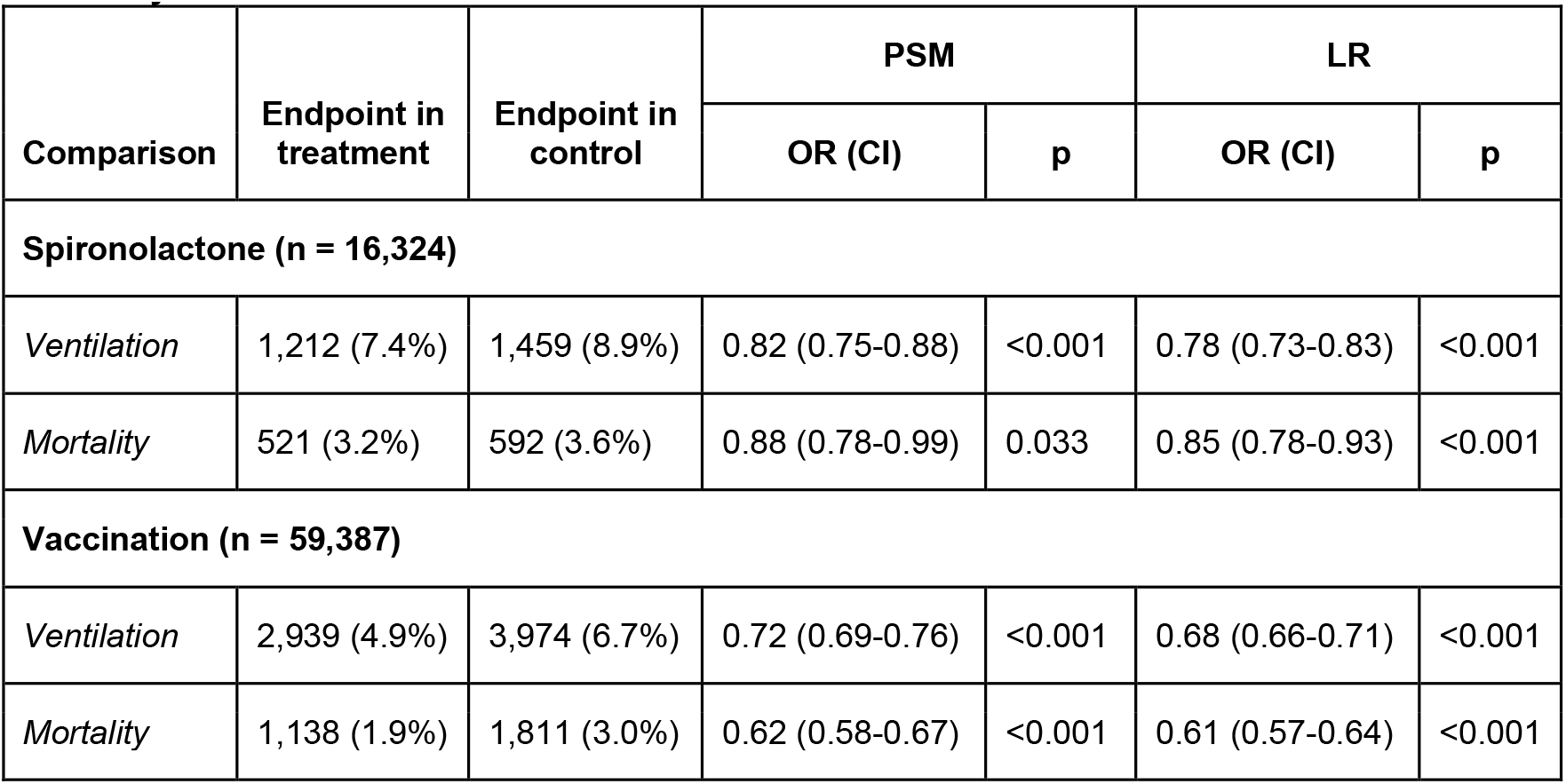
Primary outcomes

| iii. Primary Outcomes |  |  |  |  |  |  |
| --- | --- | --- | --- | --- | --- | --- |
| Comparison | Endpoint in treatment | Endpoint in control | PSM |  | LR |  |
|  |  |  | OR (CI) | p | OR (CI) | p |
| Spironolactone (n = 16,324) |  |  |  |  |  |  |
| Ventilation | 1,212 (7.4%) | 1,459 (8.9%) | 0.82 (0.75-0.88) | <0.001 | 0.78 (0.73-0.83) | <0.001 |
| Mortality | 521 (3.2%) | 592 (3.6%) | 0.88 (0.78-0.99) | 0.033 | 0.85 (0.78-0.93) | <0.001 |
| Vaccination (n = 59,387) |  |  |  |  |  |  |
| Ventilation | 2,939 (4.9%) | 3,974 (6.7%) | 0.72 (0.69-0.76) | <0.001 | 0.68 (0.66-0.71) | <0.001 |
| Mortality | 1,138 (1.9%) | 1,811 (3.0%) | 0.62 (0.58-0.67) | <0.001 | 0.61 (0.57-0.64) | <0.001 |

Spironolactone treatment was also associated with a protective effect for mortality, with 521 of 16,324 patients (3.2%) in the treatment group and 592 of 16,324 patients (3.6%) in the matched control group. In the paired analysis, this corresponded to an OR of 0.88 (95% CI: 0.78-0.99; p = 0.033), which was supported by similar findings in the regression analysis (OR 0.85; 95% CI: 0.78-0.93; p < 0.001).

As a study-level control, we also measured the effect of vaccination on both endpoints, finding strongly protective effects in all cases. For ventilation, the OR for vaccination was 0.72 (95% CI: 0.69-0.76; p < 0.001) in the paired analysis and 0.68 (95% CI: 0.66-0.71; p < 0.001) in the regression analysis. For mortality, vaccination was associated with OR values of 0.62 (95% CI: 0.58-0.67; p < 0.001) and 0.61 (95% CI: 0.57-0.64; p < 0.001) in the paired and regression analyses, respectively. These findings are consistent with previous estimates of the protective effect of vaccination in hospitalized COVID-19 patients.^19,20^

We next analyzed treatment effects in predefined patient subgroups (**Table III**). For the ventilation endpoint, we observed a stronger protective treatment effect in men than in the general population, with an OR of 0.76 (95% CI: 0.67-0.85; p < 0.001) in the matched analysis and 0.72 (95% CI: 0.66-0.79; p < 0.001) in the regression analysis.

**Table III.** Ventilation outcomes in patient subgroups

| Subgroup | No. of pairs | Endpoint in treatment | Endpoint in control | PSM |  | LR |  |
| --- | --- | --- | --- | --- | --- | --- | --- |
|  |  |  |  | OR (CI) | p | OR (CI) | p |
| **15-59** | 4,106 | 350 (8.5%) | 435 (10.6%) | 0.78 (0.67-0.91) | 0.002 | 0.73 (0.65-0.82) | <0.001 |
| **60-74** | 6,813 | 656 (9.6%) | 803 (11.8%) | 0.80 (0.71-0.89) | <0.001 | 0.79 (0.73-0.87) | <0.001 |
| **75-** | 5,405 | 206 (3.8%) | 253 (4.7%) | 0.81 (0.67-0.97) | 0.028 | 0.75 (0.65-0.87) | <0.001 |
| **Male** | 7,141 | 583 (8.2%) | 750 (10.5%) | 0.76 (0.67-0.85) | <0.001 | 0.72 (0.66-0.79) | <0.001 |
| **Female** | 9,183 | 629 (6.8%) | 718 (7.8%) | 0.87 (0.77-0.97) | 0.012 | 0.84 (0.77-0.92) | <0.001 |
| **BMI $\geq 30$** | 3,486 | 447 (12.8%) | 474 (13.6%) | 0.93 (0.81-1.07) | 0.352 | 0.89 (0.80-0.99) | 0.026 |
| **BMI <30** | 12,838 | 765 (6.0%) | 933 (7.3%) | 0.81 (0.73-0.89) | <0.001 | 0.74 (0.69-0.80) | <0.001 |
| **Delta wave** | 3,881 | 355 (9.1%) | 408 (10.5%) | 0.85 (0.73-0.99) | 0.044 | 0.76 (0.68-0.86) | <0.001 |
| **Omicron wave** | 3,329 | 203 (6.1%) | 222 (6.7%) | 0.91 (0.74-1.11) | 0.37 | 0.80 (0.69-0.93) | 0.004 |

The effect in women was weaker than in the general population, with estimated OR values of 0.87 (95% CI: 0.77-0.97; p = 0.012) and 0.84 (95% CI: 0.77-0.92; p < 0.001) in the matched and regression analyses, respectively. In non-high-BMI patients, treatment was associated with a more protective effect than in the general population, with an OR of 0.81 (95% CI: 0.73-0.89; p < 0.001) in the matched analysis and 0.74 (95% CI: 0.69-0.80) in the regression analysis. The estimated treatment effect was greatly reduced in high-BMI patients and did not meet significance in the matched analysis (OR 0.93; 95% CI: 0.81-1.07; p = 0.352), although nominally significant in the regression analysis (OR 0.89; 95% CI: 0.80-0.99; p = 0.026). Treatment was associated with a protective effect in all age brackets. However, it was most protective in the youngest bracket (<60 years old) and least protective in the oldest bracket (≥75 years old), with estimated OR of 0.78 (95% CI: 0.67-0.91; p = 0.002) and 0.81 (95% CI: 0.67-0.97; p = 0.028), respectively, in the paired analysis. The protective effect was also diminished for hospitalizations during the Omicron wave, with an OR of 0.91 (95% CI: 0.74-1.11; p = 0.37) in the paired analysis and 0.80 (95% CI: 0.69-0.93; p = 0.004) in the regression analysis.

The lower frequency of events for the mortality endpoint limited our ability to detect significant effects in subgroups for this endpoint. In the subgroup analysis for mortality (**Supplementary Table S3**), only males (OR 0.83; 95% CI: 0.70-0.98; p = 0.029) and the 60-74 age bracket (OR 0.80; 95% CI: 0.67-0.96; p = 0.017) met significance in the PSM analysis.

We also conducted sensitivity analyses to assess the effect of parameter selection in our analysis (**Table IV**). For the ventilation endpoint, treatment remained significantly associated with improved outcomes in all sensitivity analyses, including using a 90-day window (OR 0.82; 95% CI: 0.75-0.90; p < 0.001) and a 360-day window (OR 0.89; 95% CI: 0.83-0.95; p = 0.001) for drug exposure. For the mortality endpoint, significantly protective treatment effects were likewise observed for both the 90-day (OR 0.81; 95% CI: 0.70-0.93; p = 0.002) and 360-day (OR 0.85; 95% CI: 0.76-0.94; p = 0.003) windows.

**Table IV.** Sensitivity analyses

| Comparison | No. of pairs | Endpoint in treatment | Endpoint in control | PSM |  | LR |  |
| --- | --- | --- | --- | --- | --- | --- | --- |
|  |  |  |  | OR (CI) | p | OR (CI) | p |
| Ventilation |  |  |  |  |  |  |  |
| 90-day exposure window | 12,504 | 936 (7.5%) | 1,118 (8.9%) | 0.82 (0.75-0.90) | <0.001 | 0.79 (0.74-0.85) | <0.001 |
| 360-day exposure window | 20,690 | 1,524 (7.4%) | 1,701 (8.2%) | 0.89 (0.83-0.95) | 0.001 | 0.79 (0.74-0.83) | <0.001 |
| Mortality |  |  |  |  |  |  |  |
| 90-day exposure window | 12,504 | 390 (3.1%) | 479 (3.8%) | 0.81 (0.70-0.93) | 0.002 | 0.83 (0.75-0.92) | <0.001 |
| 360-day exposure window | 20,690 | 671 (3.2%) | 783 (3.8%) | 0.85 (0.76-0.94) | 0.003 | 0.87 (0.80-0.94) | <0.001 |

## DISCUSSION

Our results, supported by the largest cohort study of spironolactone in COVID-19 to date, suggest that spironolactone may improve outcomes in patients hospitalized with COVID-19. In our study, spironolactone use was associated with an 18% reduction in odds of ventilation following admission for COVID-19. This effect was more pronounced in men and in younger patients (15-59 years old), where the effects corresponded to a 22% and 24% reduction in ventilation odds, respectively. In contrast, high BMI (≥30 kg/m^2^) diminished the observed treatment effect. Spironolactone use was also associated with a significant 12% reduction in odds of mortality. In our analysis, the protective effect of spironolactone amounted to 64% and 32% of the protective effect of COVID-19 vaccination against ventilation and mortality, respectively.

Several previous studies have investigated a possible protective effect for spironolactone in COVID-19, with encouraging but underpowered results. The strongest evidence, prior to our study, was a randomized controlled trial of spironolactone-sitagliptin combination therapy in 263 patients, which demonstrated a significant improvement in a subjective clinical progression score.^12^ While rates of mortality, intensive care unit admission, intubation, and end-organ damage were reduced in this trial, the effect did not meet statistical significance. A non-randomized trial of another combination therapy, spironolactone with bromhexine, reported faster time to temperature normalization and hospital discharge in the treatment group.^11^ The largest study prior to this work was a case-control study of 6,462 patients that identified an 80% reduction in odds of spironolactone exposure in COVID-19 patients compared to matched controls, although this study was restricted to patients with liver cirrhosis.^10^

Spironolactone’s dual role as a RAAS modulator and androgen antagonist provides several plausible mechanisms for inhibition of viral entry. While clinical investigations have not demonstrated a clear relationship between RAAS modulators, such as ACE inhibitors and angiotensin II receptor blockers (ARBs), and clinical outcomes in COVID-19, there is extensive evidence that androgen signaling plays a meaningful role in viral entry.^9,21–23^ For instance, anti-androgenic drugs have been associated with protective effects in observational studies, and androgen antagonism inhibits SARS-CoV-2 cellular entry in vitro.^22,24,25^ Intriguingly, our study noted a consistent relationship between effect size and androgen levels in patient subgroups, consistent with a protective effect of spironolactone mediated by inhibition of androgen signaling. For instance, the observed effects were stronger in male, non-obese, and younger patients, all of whom tend to have higher androgen levels than their demographic counterparts.^26–28^ We also observed a smaller protective effect in hospitalizations during the predominant time period of the Omicron variant, which has been observed to rely less on androgen-dependent pathways for cellular entry.^29^ While we did not have access to laboratory data to measure a potential relationship between spironolactone effect and androgen levels directly, our results are consistent with such an association.

Our study has several limitations. Although we employ well-characterized causal inference methods like propensity score matching, the observational nature of the study precludes direct causal reasoning. Furthermore, while we control for a large collection of medical and pharmacologic covariates, unmodeled confounders may still exist that could limit the generalizability of our results. Furthermore, claims data only captures information regarding prescription issue and fulfillment and does not guarantee adherence to treatment in our control group. Claims data may also be more vulnerable to incomplete or changing coding practices than institutional medical records data. We were also unable to measure dose response in our dataset, although spironolactone dose variability is generally low.^30^

In conclusion, we show that spironolactone use is associated with improved outcomes following COVID-19 hospitalization in a nationwide cohort of nearly a million hospitalized patients. Treatment was associated with lower odds of ventilation and mortality compared to matched and unmatched controls. Furthermore, the protective effect on ventilation in patient subgroups was consistent with an androgen-dependent mechanism. Our findings support the initiation of well-powered randomized controlled trials to determine the clinical value of spironolactone in the treatment of COVID-19.

## Supporting information

STROBE Checklist

Komodo Data Disclaimer

## Data Availability

The underlying data is not publicly available due to licensing restrictions. The underlying data may be made available on reasonable request and with permission of Komodo Health.

## AUTHOR CONTRIBUTIONS

Both HCC and RBA contributed to all aspects of the manuscript (conceptualization, data acquisition, analysis, funding acquisition, investigation, methodology, project administration, resources, software, supervision, validation, visualization, drafting the manuscript, and reviewing the manuscript). Both HCC and RBA directly accessed and verified the underlying data.

