## Supplemental Tables for "Association between spironolactone use and COVID-19 outcomes in population-scale claims data: a retrospective cohort study"

**Supplementary Material**

p.2. Supplementary Table S1. Coding definitions

p.3. Supplementary Table S2. Complete list of model covariates and balance

p.5. Supplementary Table S5. Mortality outcomes in patient subgroups

Supplementary Table S1. **Coding definitions**

| **Variable** | **Associated codes** |
| --- | --- |
| Myocardial infarction | I21, I22, I252 (ICD-10-CM) |
| Congestive heart failure | I099, I110, I130, I132, I255, I420, I425, I426, I427, I428, I429, I43, I50, P290 (ICD-10-CM) |
| Peripheral vascular disease | I70, I71, I731, I738, I739, I771, I790, I792, K551, K558, K559, Z958, Z959 (ICD-10-CM) |
| Cerebrovascular disease | G45, G46, H340, I60, I61, I62, I63, I64, I65, I66, I67, I68, I69 (ICD-10-CM) |
| Dementia | F00, F01, F02, F03, F051, G30, G311 (ICD-10-CM) |
| Chronic pulmonary disease | I278, I279, J40, J41, J42, J43, J44, J45, J46, J47, J60, J61, J62, J63, J64, J65, J66, J67, J684, J701, J703 (ICD-10-CM) |
| Rheumatic disease | M05, M06, M315, M32, M33, M34, M351, M353, M360 (ICD-10-CM) |
| Peptic ulcer disease | K25, K26, K27, K28 (ICD-10-CM) |
| Mild liver disease | B18, K700, K701, K702, K703, K709, K713, K714, K715, K717, K73, K74, K760, K762, K763, K764, K768, K769, Z944 (ICD-10-CM) |
| Diabetes without chronic complications | E100, E101, E106, E108, E109, E110, E111, E116, E118, E119, E120, E121, E126, E128, E129, E130, E131, E136, E138, E139, E140, E141, E146, E148, E149 (ICD-10-CM) |
| Diabetes with chronic complications | E102, E103, E104, E105, E107, E112, E113, E114, E115, E117, E122, E123, E124, E125, E127, E132, E133, E134, E135, E137, E142, E143, E144, E145, E147 (ICD-10-CM) |
| Hemiplegia or paraplegia | G041, G114, G801, G802, G81, G82, G830, G831, G832, G833, G834, G839 (ICD-10-CM) |
| Renal disease | I120, I131, N032, N033, N034, N035, N036, N037, N052, N053, N054, N055, N056, N057, N18, N19, N250, Z490, Z491, Z492, Z940, Z992 (ICD-10-CM) |
| Malignancy | C0, C1, C20, C21, C23, C24, C25, C26, C30, C31, C32, C33, C34, C37, C38, C39, C40, C41, C43, C45, C46, C47, C48, C49, C50, C51, C52, C53, C54, C55, C56, C57, C58, C6, C70, C71, C72, C73, C74, C75, C76, C81, C82, C83, C84, C85, C88, C90, C91, C92, C93, C94, C95, C96, C97 (ICD-10-CM) |
| Moderate or severe liver disease | I850, I859, I864, I982, K704, K711, K721, K729, K765, K766, K767 (ICD-10-CM) |
| Metastatic solid tumor | C77, C78, C79, C80 (ICD-10-CM) |
| HIV/AIDS | B20, B21, B22, B24 (ICD-10-CM) |

Supplementary Table S2. **Complete list of model covariates and balance**

|  | **Spironolactone-**  **exposed** | **Non-spironolactone-exposed** | |  |
| --- | --- | --- | --- | --- |
|  |  | **Before matching** | **After matching** | **Standardized bias** |
|  | **(n = 16,324)** | **(n = 881,979)** | **(n = 16,324)** |  |
| **Age** | 67.7 (60.0-78.9) | 64.9 (56.4-78.6) | 67.3 (60.0-78.5) | 0.005 |
| *15-59* | 4,106 (25.2%) | 270,336 (30.7%) | 4,078 (25.0%) |  |
| *60-74* | 6,813 (41.7%) | 335,903 (38.1%) | 6,921 (42.4%) |  |
| *75-* | 5,405 (33.1%) | 275,740 (31.3%) | 5,325 (32.6%) |  |
| **Gender** |  |  |  | -0.028 |
| *Female* | 9,183 (56.3%) | 455,941 (51.7%) | 9,408 (57.6%) |  |
| *Male* | 7,141 (43.7%) | 426,038 (48.3%) | 6,916 (42.4%) |  |
| **BMI ≥30** | 3,486 (21.4%) | 136,505 (15.5%) | 3,490 (21.4%) | -0.001 |
| **Myocardial infarction** | 1,581 (9.7%) | 44,696 (5.1%) | 1,536 (9.4%) | 0.009 |
| **Congestive heart failure** | 7,430 (45.5%) | 121,074 (13.7%) | 7,130 (43.7%) | 0.037 |
| **Peripheral vascular disease** | 1,420 (8.7%) | 39,318 (4.5%) | 1,384 (8.5%) | 0.008 |
| **Cerebrovascular disease** | 892 (5.5%) | 54,473 (6.2%) | 911 (5.6%) | -0.005 |
| **Dementia** | 1,235 (7.6%) | 106,037 (12.0%) | 1,192 (7.3%) | 0.010 |
| **Chronic pulmonary disease** | 5,014 (30.7%) | 177,629 (20.1%) | 4,954 (30.3%) | 0.008 |
| **Rheumatic disease** | 572 (3.5%) | 20,584 (2.3%) | 595 (3.6%) | -0.008 |
| **Peptic ulcer disease** | 91 (0.6%) | 4,286 (0.5%) | 66 (0.4%) | 0.021 |
| **Mild liver disease** | 1,453 (8.9%) | 24,107 (2.7%) | 1,287 (7.9%) | 0.036 |
| **Diabetes without chronic complications** | 4,903 (30.0%) | 214,003 (24.3%) | 5,012 (30.7%) | -0.015 |
| **Diabetes with chronic complications** | 3,084 (18.9%) | 90,613 (10.3%) | 3,018 (18.5%) | 0.010 |
| **Hemiplegia or paraplegia** | 133 (0.8%) | 8,703 (1.0%) | 132 (0.8%) | 0.001 |
| **Renal disease** | 4,095 (25.1%) | 127,815 (14.5%) | 3,961 (24.3%) | 0.019 |
| **Malignancy** | 618 (3.8%) | 30,726 (3.5%) | 606 (3.7%) | 0.004 |
| **Moderate or severe liver disease** | 544 (3.3%) | 2,843 (0.3%) | 368 (2.3%) | 0.060 |
| **Metastatic solid tumor** | 114 (0.7%) | 5,918 (0.7%) | 110 (0.7%) | 0.003 |
| **HIV/AIDS** | 35 (0.2%) | 1,692 (0.2%) | 35 (0.2%) | 0.000 |
| **COVID-19 vaccination** | 1,749 (10.7%) | 57,638 (6.5%) | 1,775 (10.9%) | -0.005 |
| **Atorvastatin user** | 5,956 (36.5%) | 103,043 (11.7%) | 6,245 (38.3%) | -0.037 |
| **Levothyroxine user** | 3,505 (21.5%) | 65,560 (7.4%) | 3,624 (22.2%) | -0.018 |
| **Metformin user** | 3,405 (20.9%) | 69,896 (7.9%) | 3,721 (22.8%) | -0.048 |
| **Lisinopril user** | 3,284 (20.1%) | 70,110 (7.9%) | 3,578 (21.9%) | -0.045 |
| **Amlodipine user** | 3,367 (20.6%) | 79,004 (9.0%) | 3,758 (23.0%) | -0.059 |
| **Metoprolol user** | 5,351 (32.8%) | 77,605 (8.8%) | 5,461 (33.5%) | -0.014 |
| **Albuterol user** | 3,906 (23.9%) | 86,337 (9.8%) | 4,014 (24.6%) | -0.016 |
| **Omeprazole user** | 2,875 (17.6%) | 54,551 (6.2%) | 2,997 (18.4%) | -0.020 |
| **Losartan user** | 2,959 (18.1%) | 50,323 (5.7%) | 3,090 (18.9%) | -0.021 |
| **Gabapentin user** | 3,571 (21.9%) | 59,002 (6.7%) | 3,670 (22.5%) | -0.015 |

Supplementary Table S3. **Mortality outcomes in patient subgroups**

| **Subgroup** | **No. of pairs** | **Endpoint in treatment** | **Endpoint in control** | **PSM** | | **LR** | |
| --- | --- | --- | --- | --- | --- | --- | --- |
|  |  |  |  | **OR (CI)** | **p** | **OR (CI)** | **p** |
| **15-59** | 4,106 | 69 (1.7%) | 70 (1.7%) | 0.99 (0.70-1.38) | 1.000 | 0.83 (0.65-1.07) | 0.151 |
| **60-74** | 6,813 | 227 (3.3%) | 281 (4.1%) | 0.80 (0.67-0.96) | 0.017 | 0.81 (0.70-0.93) | 0.002 |
| **75-** | 5,405 | 225 (4.2%) | 263 (4.9%) | 0.85 (0.71-1.02) | 0.090 | 0.90 (0.78-1.03) | 0.137 |
| **Male** | 7,141 | 265 (3.7%) | 318 (4.5%) | 0.83 (0.70-0.98) | 0.029 | 0.85 (0.75-0.97) | 0.013 |
| **Female** | 9,183 | 256 (2.8%) | 280 (3.0%) | 0.91 (0.77-1.08) | 0.314 | 0.87 (0.76-0.98) | 0.028 |
| **BMI ≥30** | 3,486 | 103 (3.0%) | 114 (3.3%) | 0.90 (0.69-1.18) | 0.497 | 0.78 (0.63-0.95) | 0.016 |
| **BMI <30** | 12,838 | 418 (3.3%) | 455 (3.5%) | 0.92 (0.80-1.05) | 0.215 | 0.88 (0.80-0.98) | 0.014 |
| **Delta wave** | 3,881 | 129 (3.3%) | 138 (3.6%) | 0.93 (0.72-1.19) | 0.61 | 0.85 (0.70-1.02) | 0.072 |
| **Omicron wave** | 3,329 | 83 (2.5%) | 83 (2.5%) | 1.00 (0.73-1.36) | 1.0 | 1.01 (0.81-1.27) | 0.92 |
