## Supplementary material for "Association between spironolactone use and COVID-19 outcomes in population-scale claims data: a retrospective cohort study": Komodo Data Disclaimer

**Disclaimer regarding the use of data from Komodo Health:**

“Komodo Health, Inc. makes no representation or warranty as to the accuracy or completeness of the data (“Komodo Materials”) set forth herein and shall have, and accept, no liability or any kind, whether in contract, tort (including negligence) or otherwise, to any third party arising from or related to use of the Komodo Materials by the authors. Any use which the authors or a third party makes of the Komodo Materials, or any reliance on it, or decisions to be made based on it, are the sole responsibilities of the authors and such third party. In no way shall any data appearing in the Komodo Materials amount to any form of prediction of future events or circumstances and no such reliance may be inferred or implied.”
